# Deep Learning Evaluation of Echocardiograms to Identify Occult Atrial Fibrillation

**DOI:** 10.1101/2023.04.03.23288095

**Authors:** Nathan R. Stein, Grant Duffy, Roopinder K. Sandhu, Sumeet S. Chugh, Christine M. Albert, Susan Cheng, David Ouyang, Neal Yuan

## Abstract

**Background:** Atrial fibrillation (AF) can often be missed by intermittent screening given its frequently paroxysmal and asymptomatic presentation. Deep learning algorithms have been developed to identify patients with paroxysmal AF from electrocardiograms (ECGs) in sinus rhythm. Transthoracic echocardiograms (TTEs) may provide additional structural information complementary to ECGs that could also be used to help identify occult AF.

**Objective:** We sought to determine whether deep learning evaluation of echocardiograms of patients in sinus rhythm could identify occult AF.

**Methods:** We identified patients who had TTEs performed between 2004 and 2021. We created a two-stage model that (1) distinguished which TTEs were in sinus rhythm and which were in AF and then (2) predicted which of the TTEs in sinus rhythm were in patients with paroxysmal AF. Models were trained from video-based convolutional neural networks using TTE parasternal long axis (PLAX) videos. The AF prediction performance was compared to prediction using clinical variables, CHADSVASc score, and left atrial (LA) size.

**Results:** Our model trained on 111,319 TTE videos distinguished TTEs in AF from those in sinus rhythm with high accuracy (AUC 0.96, 0.95-0.96). A total of 72,181 TTE videos were in sinus rhythm. When tested on a held-out sample, the model predicted the occurrence of concurrent AF with an AUC of 0.71 (0.69-0.73). Using the max F1 threshold, the PPV was 0.20 and the NPV was 0.95. The model performed better than predicting concurrent AF using clinical risk factors (AUC 0.67, 0.65-0.69), LA area (AUC 0.63, 0.62-0.64), and CHADSVASc (AUC 0.61, 0.60-0.62).

**Conclusion:** A deep learning model distinguished AF from sinus rhythm TTEs with high accuracy and predicted the presence of AF within 90 days of sinus rhythm TTEs moderately well, better than clinical variables or LA size alone. TTEs may help inform automated opportunistic AF screening efforts.

## INTRODUCTION

Atrial fibrillation (AF) is the most common cardiac arrhythmia^1^ and is associated with significant morbidity and mortality^2^. AF is frequently paroxysmal and asymptomatic^3^, and therefore undetected until complications such as stroke or heart failure present. Given the potential promise of intervening on early AF, with anticoagulation to reduce stroke risk or therapies to maintain early sinus rhythm, a number of trials have investigated the utility of routine screening for AF using either home or office based intermittent and/or continuous electrocardiography (ECG) ^4–13^. These studies reveal that many cases of AF are not detected with conventional screening practices.

Recent work has shown that artificial intelligence (AI) applied to ECGs can predict incident AF from sinus ECGs ^14–16^. This work has even been validated in prospective clinical trials^17^, suggesting the value and efficacy of opportunistic screening by AI. Transthoracic echocardiograms (TTEs) are routinely obtained tests in patients with cardiovascular symptoms and individuals who are at high risk for AF. The most common cardiovascular imaging modality, TTEs may provide additional structural information complementary to ECGs that could also be opportunistically used to help identify occult AF. We have previously shown that video-based AI can determine left atrial and ventricular sizes, presence of pacemaker leads,^18^ as well as determine ejection fraction with variance comparable to or less than human experts^19^. In this study, we sought to determine whether a deep learning model using echocardiogram videos could identify patients in AF, including even those in sinus rhythm at the time of the echocardiogram.

## METHODS

### Dataset

We identified all TTEs performed between 6/2004 and 6/2021 at Cedars-Sinai Medical Center. We included only those TTEs that were in AF (atrial fibrillation or atrial flutter) or sinus rhythm per the TTE report and by the absence or presence of an identified mitral A wave doppler velocity. TTEs in sinus rhythm were additionally required to have a sinus rhythm ECG within 24 hours. Among those TTEs in sinus rhythm, we further classified paroxysmal AF cases as patients who had a TTE in sinus rhythm with AF documented on ECG within 90 days before/after the TTE. Control patients had a sinus rhythm TTE and no documented AF within 90 days by ECG and no AF by ICD (International Classification of Diseases) diagnosis prior to or up to 90 days after the TTE (**Figure 1**). The 90 day interval was chosen as a cutoff, since we hypothesized that patients who were having paroxysmal AF would likely already demonstrate some structural changes on TTE that our model could pick up. We performed additional modeling with a 365 day window as well. Patient characteristics and comorbidities were derived from electronic health records using Elixhauser ICD comorbidity definitions^20^ to calculate the CHADSVASc score. The left atrial (LA) size was obtained from the TTE report.

**Figure 1.**
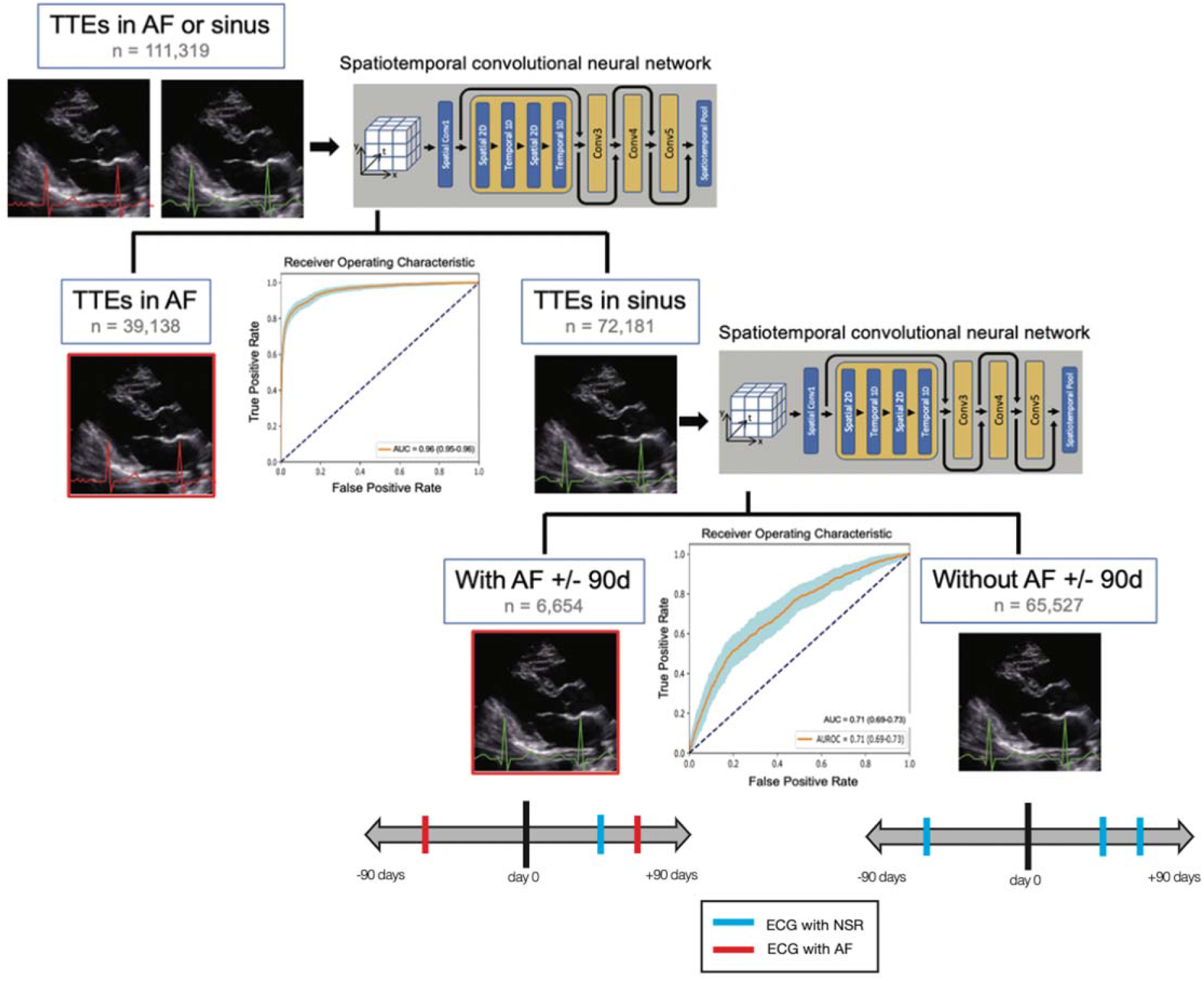
Determination of SR, AF, and concurrent AF. TTEs were first stratified by being in AF or SR. The model was able to determine AF versus SR (AUC 0.96). Of the subset in SR, cases were defined as patients who had a TTE in SR (per the TTE report and confirmed by presence of noted mitral A wave velocity) with AF documented on ECG within 90 days before/after the TTE. Controls had a SR TTE and no documented AF within 90 days by ECG and no AF by ICD (International Classification of Diseases) diagnosis prior to or up to 90 days after the TTE. The model was able to determine the presence or absence of concurrent AF (AUC 0.71).

TTEs were acquired using Philips EPIQ 7 or iE33 ultrasound machines. For deep learning, each TTE study was initially sourced in Digital Imaging and Communications in Medicine (DICOM) format and contained multiple video loops. Videos corresponding to the Parasternal Long Axis (PLAX) view were extracted, masked, and down sampled per previously described methods^19^ prior model input. The PLAX view was chosen because it is reliably present in most TTE studies and captures important information regarding left ventricular function, mitral valve disease, as well as left atrial size. For model training and validation, the TTE dataset was split using 80% of TTEs for model training, 10% for validation, and 10% for hold-out testing. The study was approved by the IRB at Cedars-Sinai Medical Center.

### Deep learning model selection and training

We trained a convolutional neural network with residual connections and spatiotemporal convolutions using the R2+1D architecture to determine whether a TTE was in sinus rhythm or AF, and subsequently to predict concurrent paroxysmal AF from TTE videos^19,21^. We initialized this model using pretrained weights from the EchoNet-Dynamic dataset^19^. Models were trained to minimize the squared loss between the predicted risk and the actual label (0 for no AF and 1 for AF) with an Adam optimizer set to a learning rate of 0.001. We used a batch size of 64 across 50 epochs. The model was fed video clips of 32 frames created by sampling every other frame. All model training was done using the Python library PyTorch.

### Model performance

Model weights from the epoch with the best validation loss were used for final model performance testing on the held out dataset. We created receiver operating characteristic (ROC) and precision-recall (PR) curves to show model performance across different classification thresholds. The overall model performance was summarized using the area under the curve (AUC) for the ROC curve as well as the positive predictive value (PPV), negative predictive value (NPV), and the F1 score at the maximum F1 threshold. We report confidence intervals using 10,000 bootstrapped samples.

We compared our deep learning model to AF prediction by logistic regression using clinical variables (age, sex, heart failure, hypertension, cerebrovascular disease, peripheral arterial disease, diabetes, height, weight, smoker), CHADSVASc score, and left atrial (LA) size on TTE. These statistical analyses were conducted using R software (version 3.4.1, Vienna, Austria).

## RESULTS

Our cohort included a total of 111,319 TTEs, of which 39,138 studies were in AF and 72,181 studies were in sinus rhythm. Among those in sinus rhythm, 6,654 studies were in patients with concurrent paroxysmal AF, while 65,527 studies were in patients without concurrent AF. Patients in atrial fibrillation were on average older (75.2 vs. 66.0 years old), less often female (40.1% vs. 45.5%), more often White (67.9% vs. 57.1%), and had more comorbidities (Table 1). The average left atrium area was larger (26.0 vs. 19.3 cm^2^) and CHADSVASc score was marginally higher (3.9 vs. 3.1). Among those in sinus rhythm, patients with concurrent paroxysmal AF were also older (72.1 vs. 65.3 years old), less often female (39.93% vs. 45.9%), more often White (64.2% vs. 56.4%), and had more comorbidities. Left atrial area was slightly larger (21.8 vs. 19.1 cm^2^) and CHADSVASc score was higher (3.8 vs. 3.0).

**Table 1.** Demographics and clinical characteristics for patients with TTEs in AF and sinus rhythm. Further stratified by patients who were in sinus rhythm at time of TTE but had concurrent AF within 90 days.

|  | AF | Sinus | p-value | Sinus With<br>Concurrent AF | Sinus Without<br>Concurrent AF | p-value |
| --- | --- | --- | --- | --- | --- | --- |
| n | 39138 | 72181 |  | 6654 | 65527 |  |
| Age | 75.2 (13.1) | 66.0 (16.9) | <0.001 | 72.1 (13.6) | 65.3 (17.1) | <0.001 |
| Female | 15694 (40.1) | 32755 (45.4) | <0.001 | 2657 (39.9) | 30098 (45.9) | <0.001 |
| Race |  |  | <0.001 |  |  | <0.001 |
| American Indian | 80 (0.2) | 155 (0.2) |  | 10 (0.2) | 145 (0.2) |  |
| Asian | 2539 (6.5) | 5100 (7.1) |  | 547 (8.2) | 4553 (7.0) |  |
| Black | 4258 (10.9) | 11746 (16.3) |  | 759 (11.4) | 10987 (16.8) |  |
| Hispanic | 2845 (7.3) | 7462 (10.4) |  | 531 (8.0) | 6931 (10.6) |  |
| White | 26525 (67.9) | 41088 (57.1) |  | 4270 (64.2) | 36818 (56.4) |  |
| Other | 2170 (5.6) | 5265 (7.3) |  | 370 (5.6) | 4895 (7.5) |  |
| Pacific Islander | 72 (0.2) | 180 (0.3) |  | 15 (0.2) | 165 (0.3) |  |
| Unknown | 552 (1.4) | 959 (1.3) |  | 145 (2.2) | 814 (1.2) |  |
| Heart Failure | 18697 (47.8) | 19574 (27.1) | <0.001 | 2865 (43.1) | 16709 (25.5) | <0.001 |
| Hypertension | 24881 (63.6) | 39101 (54.2) | <0.001 | 4163 (62.6) | 34938 (53.3) | <0.001 |
| Prior CVA | 10816 (27.6) | 18150 (25.1) | <0.001 | 2045 (30.7) | 16105 (24.6) | <0.001 |
| Prior Myocardial Infarction | 4757 (12.2) | 8443 (11.7) | 0.025 | 1155 (17.4) | 7288 (11.1) | <0.001 |
| Peripheral Artery Disease | 6378 (16.3) | 7560 (10.5) | <0.001 | 1183 (17.8) | 6377 (9.7) | <0.001 |
| Diabetes | 9391 (24.0) | 16507 (22.9) | <0.001 | 1789 (26.9) | 14718 (22.5) | <0.001 |
| Smoker | 170 (11.5) | 168.2 (11.1) | <0.001 | 273 (4.1) | 2820 (4.3) | 0.460 |
| Height (cm) | 79.73 (22.3) | 78.2 (21.1) | <0.001 | 169 (11.2) | 168.2 (11.1) | <0.001 |
| Weight (kg) | 1752 (4.5) | 3093 (4.3) | 0.139 | 78.4 (20.2) | 78.23 (21.2) | 0.631 |
| LA Area (cm<sup>2</sup>) | 26.0 (8.2) | 19.3 (5.7) | <0.001 | 21.8 (6.4) | 19.07 (5.6) | <0.001 |
| LV Ejection Fraction | 52.6 (16.1) | 59.1 (13.4) | <0.001 | 55.97 (15.4) | 59.4 (13.1) | <0.001 |
| CHADS2VASc | 3.9 (2.1) | 3.1 (2.1) | <0.001 | 3.84 (2.1) | 3.0 (2.1) | <0.001 |

When tested on a held-out dataset, our deep learning model distinguished whether a TTE was in AF or sinus rhythm with an AUC of 0.96 (0.95-0.96) (**Figure 1**). Among those TTEs in sinus rhythm, the model predicted concurrent paroxysmal AF with an AUC of 0.71 (0.69-0.73) (**Figure 2**). Using the max F1 threshold, the PPV was 0.20, the NPV was 0.95, and the F1 score was 0.29. The model performed better than predicting concurrent paroxysmal AF using clinical risk factors (AUC 0.67), LA area (AUC 0.63), and CHADSVASc score (AUC 0.61).

**Figure 2.**
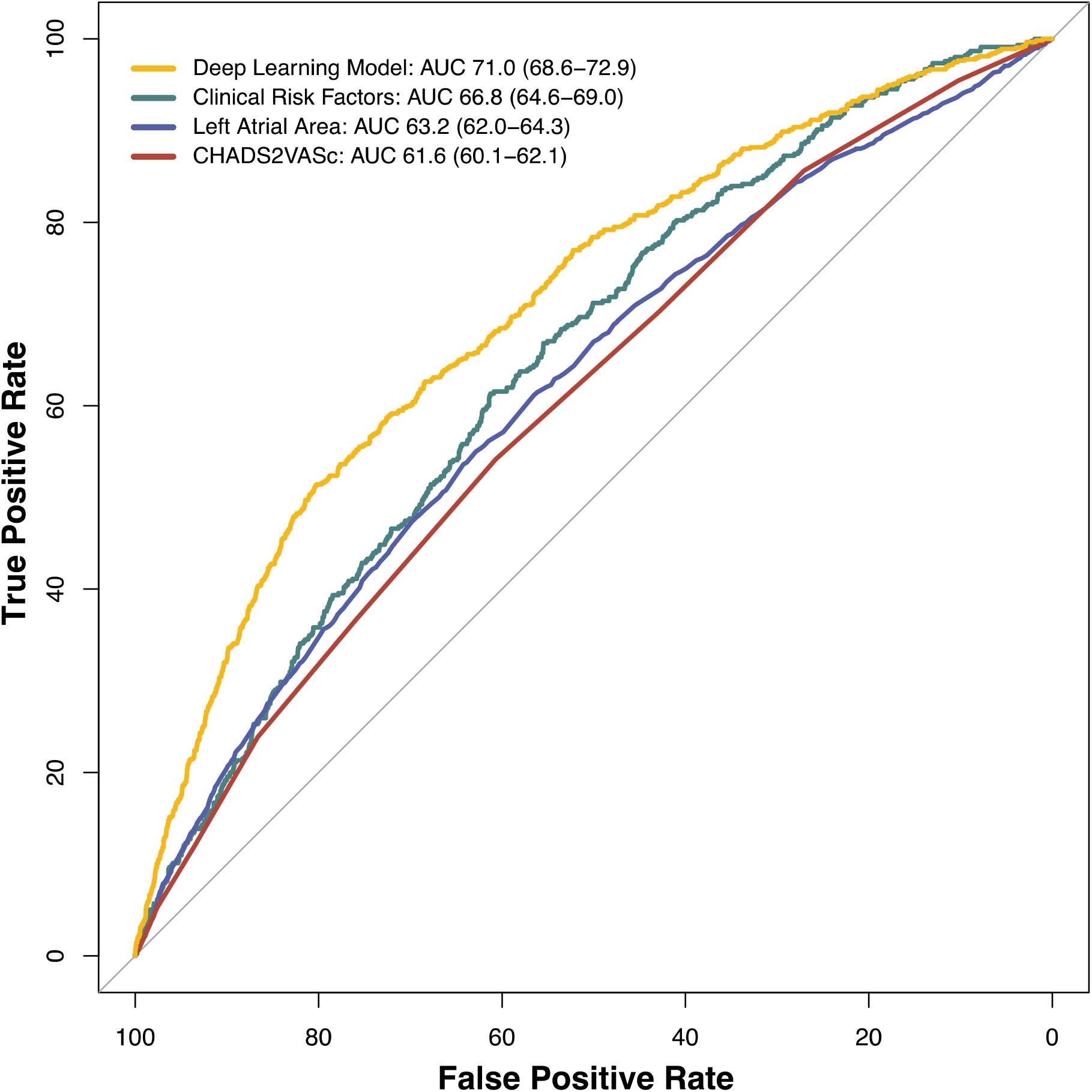
Prediction of concurrent paroxysmal atrial fibrillation. Performance of a deep learning model for prediction of concurrent paroxysmal AF from PLAX TTE videos in sinus rhythm compared to models using clinical risk factors, left atrial area, and CHADSVASc score.

## DISCUSSION

In this study, we found that a deep learning model trained on echocardiogram videos could distinguish a high risk patient group by identifying both patients in AF (without the use of rhythm or doppler data) as well as those in sinus rhythm with concurrent paroxysmal AF. The model outperformed the prediction of concurrent paroxysmal AF using CHADSVASc, left atrial area, or clinical risk factors alone. While the accuracy for predicting concurrent AF was moderate, applying such a model to all routinely acquired TTEs could present an opportunity for improving future efforts to screen for AF and prevent its complications.

AF is frequently asymptomatic^22^ and therefore goes unidentified until complications such as a stroke occur. In fact, occult AF is detected in up to 20% of patients with acute stroke^23,24^. Being able to identify patients with occult AF in patients with prior stroke can be crucial, as initiation of anticoagulation can reduce the risk of future stroke^25^. Additionally, earlier identification of occult AF can allow initiation of rhythm control strategies, which may have more beneficial long term outcomes^26–29^.

Echocardiography is a noninvasive test frequently performed in patients who are at risk for unrecognized paroxysmal AF whether for direct evaluation of the etiology of thromboembolism or stroke or for other cardiac pathologies that are correlated with high AF risk^30^. TTE videos are likely able to capture some of the structural changes in the heart that may signal ongoing atrial fibrillation, even when the heart is in sinus rhythm. Echocardiographic parameters including left atrial size and function ^31–33^, left ventricular wall thickness ^34^, diastolic function ^35^, LAVI/a’ (ratio of LA volume index to tissue Doppler A’) ^36,37^, and septal PA-TDI (atrial conduction time) ^38,39^ have been used to identify patients with AF. However, all of these assessments require specific measurements to be performed at the time of the TTE and may not be measurable in all patients. A major strength of our model is that it only requires a single PLAX clip, as oppose to a whole echocardiogram, and does not require any doppler information. The PLAX is a highly standardized and easily obtainable view, even by novice scanners, and has previously been used on deep learning models for aortic stenosis and coronary artery calcification^21,40^. By requiring only a short clip from the PLAX view, our model is an efficient and pragmatic approach and could allow this tool to be used in real-time practice.

Our model performed well in identifying active AF during the time of TTE. Though AF is readily identified by trained cardiologists, automated rhythm identification may be useful for providers performing bedside and point-of-care ultrasounds. The identification of patients with paroxysmal AF at the time of a sinus rhythm TTE is more challenging. There are currently numerous methods to predict occult AF, including using clinical variables, imaging data, and electrophysiologic data ^41^. Our model was able to identify concurrent paroxysmal AF better than prediction using the CHADSVASc score or risk factors included in other clinical AF risk scores such as CHARGE-AF ^42^. However, it performed less well than deep learning models using ECG data^15^. This suggests that structure alone is insufficient in determining concurrent AF, consistent with there being changes in the electrical system of the LA that precede the structural changes seen on imaging. Indeed, previous work has shown that analysis of P wave morphologies and dispersion^43,44^ and atrial premature complexes^45^ can predict AF. Due to the interplay between atrial electrical activity and structure, as LA enlargement can beget AF and also be caused by longstanding AF, it is possible that future deep learning analysis of simultaneous TTE and ECG data may be synergistic and lead to improved predictions.

A current application of our model may be as an additional screening test for AF in patients with prior stroke, and to guide subsequent testing. Our PPV at the maximum F1 threshold was 0.20, which would give a number needed to screen of 5 patients. Though further prospective testing is needed to validate our model, since TTE is already performed in the routine evaluation after a stroke^37^, being able to glean additional insights from this non-invasive test is promising. Current guidelines state that prolonged rhythm monitoring to screen for AF after a stroke is reasonable, though duration remains uncertain^30^. Since longer periods of ambulatory rhythm monitoring lead to increased AF detection rates^46^, a positive TTE screen could be used to guide clinical decision making for duration of screening and wearable versus implantable monitoring.

There are several limitations to our study that should be addressed. First, there could be labeling inaccuracies, where patients with concurrent paroxysmal AF were not detected by ECGs in our system or entered into the EHR by ICD code. However, this would have biased our model to the null and cause underperformance. While the prevalance of concurrent paroxysmal AF was similar to other studies at academic centers^15^, the prevalence of AF in our cohort was particularly high due to our study’s design. We required that sinus rhythm TTEs have both a TTE in SR by TTE report *and* an ECG within 24 hours showing SR, which likely reduced the number of included sinus TTEs in our cohort. This therefore increased the relative prevalance of AF TTEs. While the higher proportion of AF cases does not reflect prevalence in the general population, we believe that rigorously defining TTEs in SR increased the quality of our model training and therefore validity. Our analyses were done on data from a single center which may limit generalizability. In order to verify the performance and utility of our model, further validation on an external data set as well as prospective analyses is needed.

## CONCLUSION

A deep learning model determined whether a TTE was in SR or AF with high accuracy and was able to predict the presence of concurrent paroxysmal atrial fibrillation from sinus TTEs moderately well. The model performed better than what could be predicted from clinical variables or LA size alone. This may offer additional opportunities to guide patient screening for occult AF by identifying patients who may benefit from longer term monitoring.

## Data Availability

All data produced in the present study are available upon reasonable request to the authors.

